# Long COVID brain fog and muscle pain are associated with longer time to clearance of SARS-CoV-2 RNA from the upper respiratory tract during acute infection

**DOI:** 10.1101/2023.01.18.23284742

**Authors:** Annukka A. R. Antar, Tong Yu, Zoe O Demko, Chen Hu, Jeffrey A. Tornheim, Paul W. Blair, David L. Thomas, Yukari C. Manabe, OutSMART Study Group

**Affiliations:** Department of Medicine, Johns Hopkins University School of Medicine, Baltimore, MD, United States; Department of Oncology, Johns Hopkins University School of Medicine, Baltimore, MD, United States; Henry M. Jackson Foundation for the Advancement of Military Medicine, Inc., Bethesda, MD, United States

**Author notes:** **Correspondence:** Dr. Annukka Antar.

**Keywords:** COVID-19, Long COVID, Post-Acute COVID-19 Syndrome, Post-Acute Sequelae of SARS-CoV-2 infection, Brain fog, Cognitive Dysfunction, Pain, Reservoir

## Abstract

The incidence of long COVID is substantial, even in people who did not require hospitalization for acute COVID-19. The pathobiological mechanisms of long COVID and the role of early viral kinetics in its development are largely unknown. Seventy-three non-hospitalized adult participants were enrolled within approximately 48 hours of their first positive SARS-CoV-2 RT-PCR test, and mid-turbinate nasal and saliva samples were collected up to 9 times within the first 45 days after enrollment. Samples were assayed for SARS-CoV-2 using RT-PCR and additional test results were abstracted from the clinical record. Each participant indicated the presence and severity of 49 long-COVID symptoms at 1-, 3-, 6-, 12-, and 18-months post-COVID-19 diagnosis. Time from acute COVID-19 illness onset to SARS-CoV-2 RNA clearance greater or less than 28 days was tested for association with the presence or absence of each of 49 long COVID symptoms at 90+ days from acute COVID-19 symptom onset. Brain fog and muscle pain at 90+ days after acute COVID-19 onset were negatively associated with viral RNA clearance within 28 days of acute COVID-19 onset with adjustment for age, sex, BMI ≥ 25, and COVID vaccination status prior to COVID-19 (brain fog: aRR 0.46, 95% CI 0.22-0.95; muscle pain: aRR 0.28, 95% CI 0.08-0.94). This work indicates that at least two long COVID symptoms - brain fog and muscle pain – at 90+ days from acute COVID-19 onset are specifically associated with longer time to clearance of SARS-CoV-2 RNA from the upper respiratory tract.

## Introduction

The incidence of long COVID is substantial, even in people who did not require hospitalization for acute COVID-19 (1). The United States Census Household Pulse Survey of December 2022 estimates that 28% of U.S. adults who ever had COVID-19 experienced long COVID symptoms lasting 3 months or longer (2). Among the most common and debilitating symptoms are pain and a lack of mental clarity or problems with concentration, also called “brain fog” (3, 4). Viral antigen persistence in tissues may be one mechanism of long COVID (5, 6). However, few longitudinal studies with well-characterized viral dynamics in the acute phase have assessed the relationship between viral persistence and long COVID symptoms, especially in people with mild to moderate acute COVID-19 who represent typical disease severity in the general population. Here we examine the association between time to viral RNA clearance from the upper respiratory tract (URT) during acute COVID-19 and specific clinical symptoms of long COVID at 90 or more days from acute COVID-19 illness onset in a cohort of people diagnosed with symptomatic COVID-19 in the outpatient setting.

## Methods

A convenience sample of non-hospitalized adults with a first positive SARS-CoV-2 PCR test within the previous 48 hours from the Johns Hopkins Health System and their household contacts were prospectively enrolled between April 21, 2020, and October 28, 2021 (7). Participants provided verbal consent after documentation of understanding, using a consent waiver with an alteration of informed consent, as most were isolating or quarantining at home. The study was conducted in either English or Spanish. This study protocol and verbal consent were approved by the Johns Hopkins University School of Medicine Institutional Review Board.

Mid-turbinate nasal swabs and saliva (URT samples) were collected at 1, 3, 7, 10, 14, 21, 28, 35, and 42 days from enrollment. Mid-turbinate nasal swabs were collected in 600 μL volumes of viral transport media, and 200μL volumes of saliva were collected. Collections at 1, 3, 7, 10, 14, 21, 35, and 42 days after study enrollment were self-collected into study-provided vials with guidance by phone or video call from study staff and frozen immediately. Collections at 28 days after study enrollment were performed primarily in-person at a research clinic visit with study staff guiding or performing collection. SARS-CoV-2 RT-PCR testing was performed on all samples (Abbott *m*2000 platform, Abbott Park, IL, USA) per manufacturer instructions as described previously (8). Discordant results from samples collected on the same day (e.g. positive nasal, negative saliva) were recorded as URT positive. Adequacy of self-collected samples was confirmed via RT-PCR for gapDH human gene expression utilizing a TaqMan gene expression assay for the first 1338 samples (8). Briefly, eluates from the Abbott *m*2000 were analyzed by the NanoDrop One (ThermoFisher Scientific, Waltham, MA) to determine total RNA concentration and all samples were normalized to 1 ng/μL of total RNA prior to analysis for gapDH. Normalized samples were utilized to synthesize cDNA in 10μL volumes utilizing Superscript IV RT (Thermofisher) per manufacturer instructions. Post cDNA synthesis, samples were assessed for the presence of gapDH utilizing RT-PCR (Applied Biosystems, Foster City, CA) per manufacturer instructions. Positive reactions were defined as reactions having a cycle threshold value < 40. 1227 of 1338 self-collected samples were positive for human gapDH, and all negative reactions started from eluates with < 1 μL, indicating insufficient material for NanoDrop analysis or cDNA synthesis. COVID-19 PCR testing results from the clinical electronic medical records were also included in this analysis.

At 1, 3, 6, 12, and 18 months post-COVID-19 diagnosis, participants were asked, “Have you had any of the following symptoms in the past week?” and selected one of “Not at all”, “A little bit”, “Somewhat”, “Very much”, or “Quite a bit” for each of 49 long COVID symptoms (Table 1). Thirty-eight symptoms derive from the FLU-PRO© Plus (9), and eleven additional symptoms were added based on frequency of report in a Long COVID Patient-Led Research Collaborative survey with input from the patient-researcher authors (3). Similar to the other symptoms, brain fog was assessed with the question “Have you had problems with concentration or ‘brain fog’ in the past week?”

**TABLE 1.**
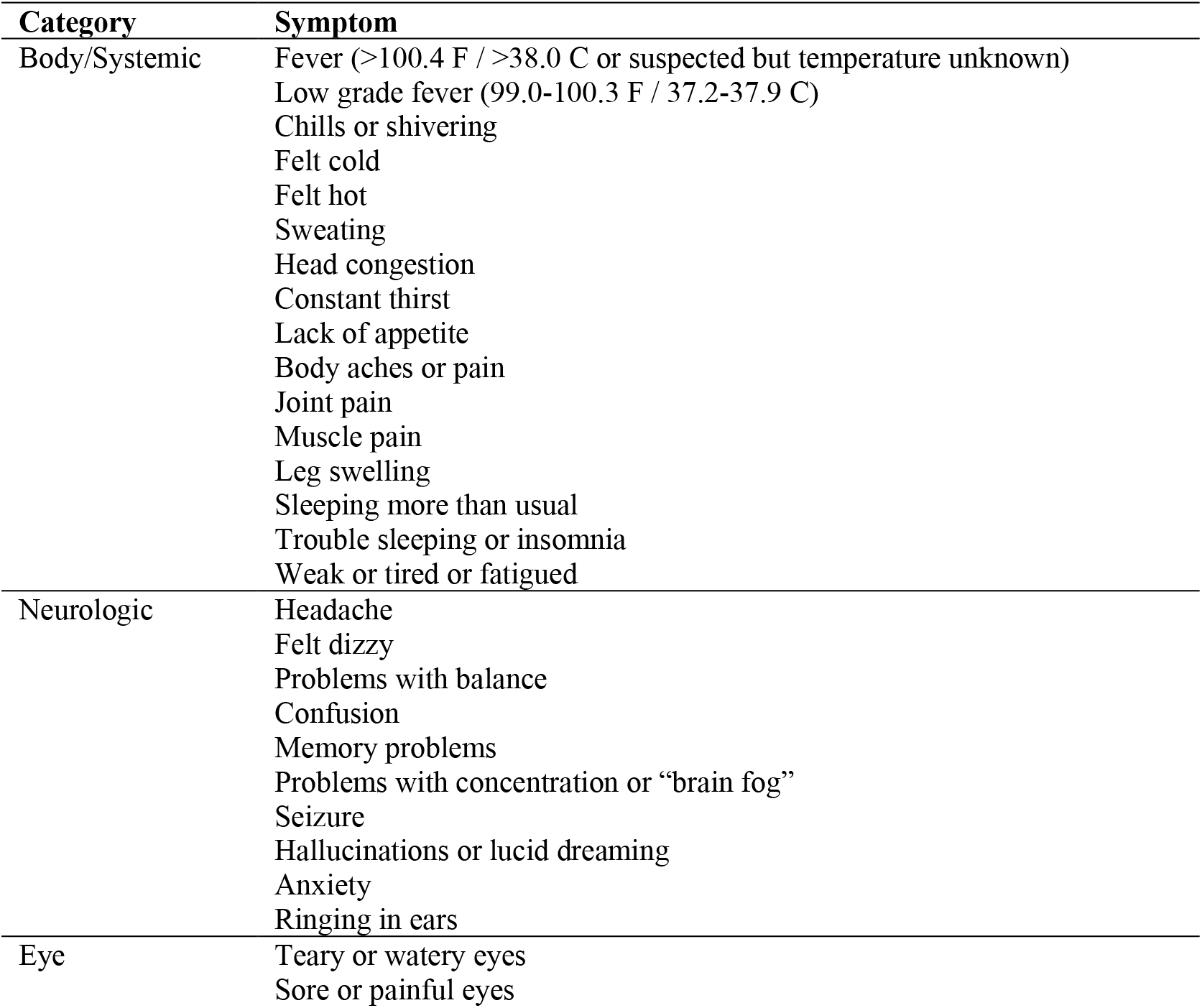
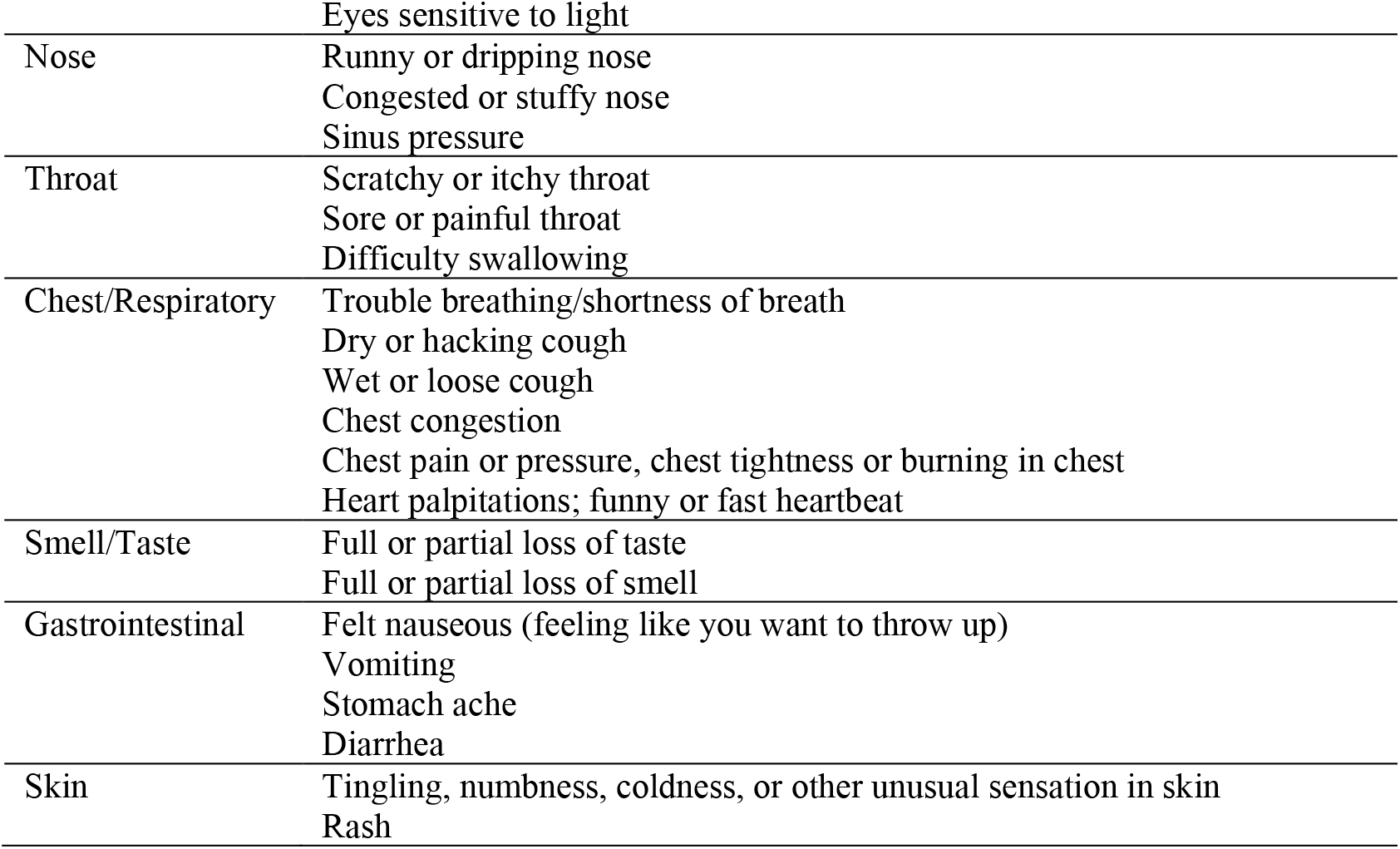
List of 49 long COVID symptoms assessed in surveys 1, 3, 6, 12, and 18 months from enrollment. Participants were asked, “Have you had any of the following symptoms in the past week?” and selected one of “Not at all”, “A little bit”, “Somewhat”, “Very much”, or “Quite a bit” for each symptom.

Time to SARS-CoV-2 RNA clearance was defined as the number of days between acute COVID-19 symptom/illness onset (recorded at study enrollment) and the midpoint between a participant’s last positive URT RNA test and the following negative URT PCR test. Time to viral RNA clearance was classified as a binary variable: within or beyond 28 days. For each of 49 long COVID symptoms, we estimated the relative risk of reporting that symptom 90+ days from acute COVID-19 onset by time to viral RNA clearance within 28 days using log-binomial regression (10) and adjusting for age, sex, BMI over/under 25, and COVID vaccination status prior to acute CVOID-19 (1+ vaccines vs. none) (StataSE 17, StataCorp).

## Results

Seventy-three participants with symptomatic, PCR-confirmed acute COVID-19, age ≥ 18, with ≥ 2 URT samples within 28 days of enrollment, and who completed at least one comprehensive symptom survey ≥ 90 days from acute COVID-19 onset were included in this analysis. The participants had a median of three study days with a positive PCR test prior to a study day with a negative PCR test, and 75% of participants had ≥ 5 study days with PCR tests available for analysis. The median time to viral RNA clearance was 17 days for the 51 participants who had a negative PCR test within 21 days of their last positive PCR test (interquartile ratio, IQR, 14.75-22.75).

The median age of participants was 52 years (IQR, 42-60), 63% were women, 27% self-reported as non-Hispanic African American, 52% self-reported as non-Hispanic White, and 16% self-reported as Hispanic (Table 2). All participants tested positive for COVID-19 in the outpatient setting, and two (2.7%) ultimately required hospitalization during acute COVID-19. Of the participants, 81% were unvaccinated prior to COVID-19, 5% had incomplete primary vaccine series, and 14% had received a complete primary vaccine series prior to COVID-19 (Table 2).

**TABLE 2.**
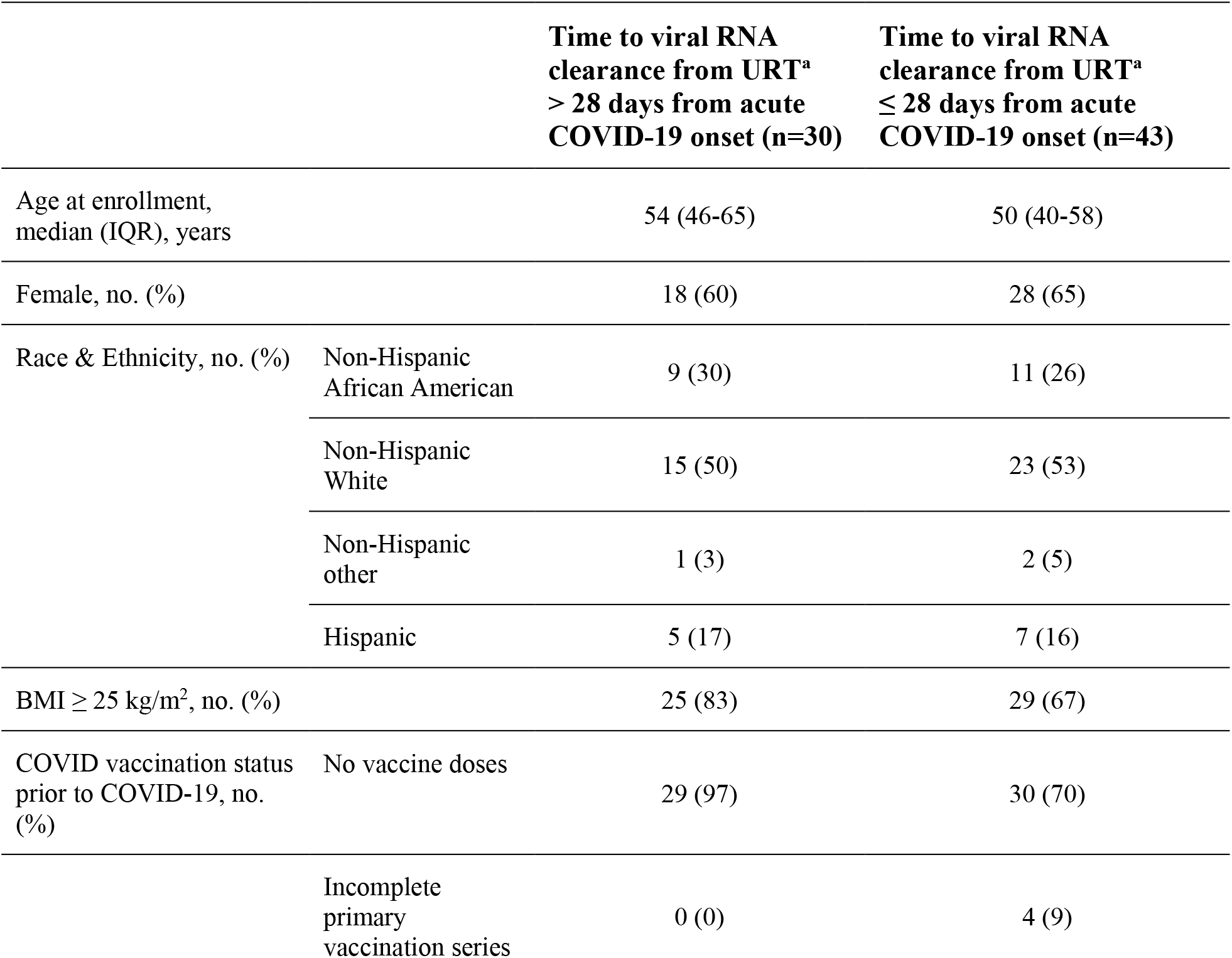
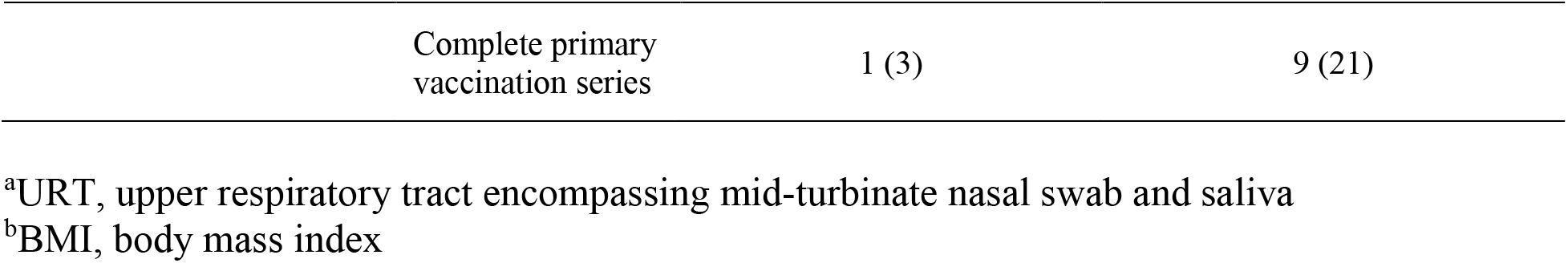
Participant demographics, body mass index, and pre-COVID vaccination statustab.

|  |  | **Time to viral RNA**<br>**clearance from URT<sup>a</sup>**<br>**> 28 days from acute**<br>**COVID-19 onset (n=30)** | **Time to viral RNA**<br>**clearance from URT<sup>a</sup>**<br>**≤ 28 days from acute**<br>**COVID-19 onset (n=43)** |
| --- | --- | --- | --- |
| Age at enrollment,<br>median (IQR), years |  | 54 (46-65) | 50 (40-58) |
| Female, no. (%) |  | 18 (60) | 28 (65) |
| Race & Ethnicity, no. (%) | Non-Hispanic<br>African American | 9 (30) | 11 (26) |
|  | Non-Hispanic<br>White | 15 (50) | 23 (53) |
|  | Non-Hispanic<br>other | 1 (3) | 2 (5) |
|  | Hispanic | 5 (17) | 7 (16) |
| BMI ≥ 25 kg/m<sup>2</sup>, no. (%) |  | 25 (83) | 29 (67) |
| COVID vaccination status<br>prior to COVID-19, no.<br>(%) | No vaccine doses | 29 (97) | 30 (70) |
|  | Incomplete<br>primary<br>vaccination series | 0 (0) | 4 (9) |

|  |  |  |
| --- | --- | --- |
| Complete primary<br>vaccination series | 1 (3) | 9 (21) |

<sup>a</sup>URT, upper respiratory tract encompassing mid-turbinate nasal swab and saliva
<sup>b</sup>BMI, body mass index

The association of the presence of each of 49 long COVID symptoms (Table 1) at 90+ days post-acute COVID-19 onset with time to SARS-CoV-2 RNA clearance from the URT within 28 days was first tested without adjustment using log-binomial regression analyses. Eleven symptoms with p values < 0.2 were tested using log-binomial regression analyses with adjustment for age, sex, BMI ≥ 25, and COVID vaccination status prior to acute COVID-19. Brain fog and muscle pain at 90+ days after acute COVID-19 illness onset were negatively associated with viral RNA clearance within 28 days with these adjustments (Figure, brain fog: aRR 0.46, 95% CI 0.22-0.95; muscle pain: aRR 0.28, 95% CI 0.08-0.94). The model failed to converge for two rarely reported symptoms, leg swelling and problems with balance. Leg swelling at 90+ days from acute COVID-19 onset was negatively associated with viral RNA clearance within 28 days of acute COVID-19 onset in an unadjusted analysis (RR 0.17, 95% CI 0.04-0.76) and in an analysis adjusted only for age and sex (aRR 0.21, 95% CI 0.05-0.82). Problems with balance at 90+ days from acute COVID-19 onset was negatively associated with viral RNA clearance within 28 days of acute COVID-19 onset only in an analysis adjusted for age, sex, and vaccination status (aRR 0.47, 95% CI 0.25-0.90) but not in a univariate analysis. There were nonsignificant trends towards negative associations of body aches or pain, anxiety, confusion, and sweating with viral RNA clearance within 28 days after adjustment for age, sex, BMI ≥ 25, and COVID vaccination status prior to acute COVID-19 (Figure). No other associations of long COVID symptoms with viral RNA clearance time within 28 days reached statistical significance.

**FIGURE.**
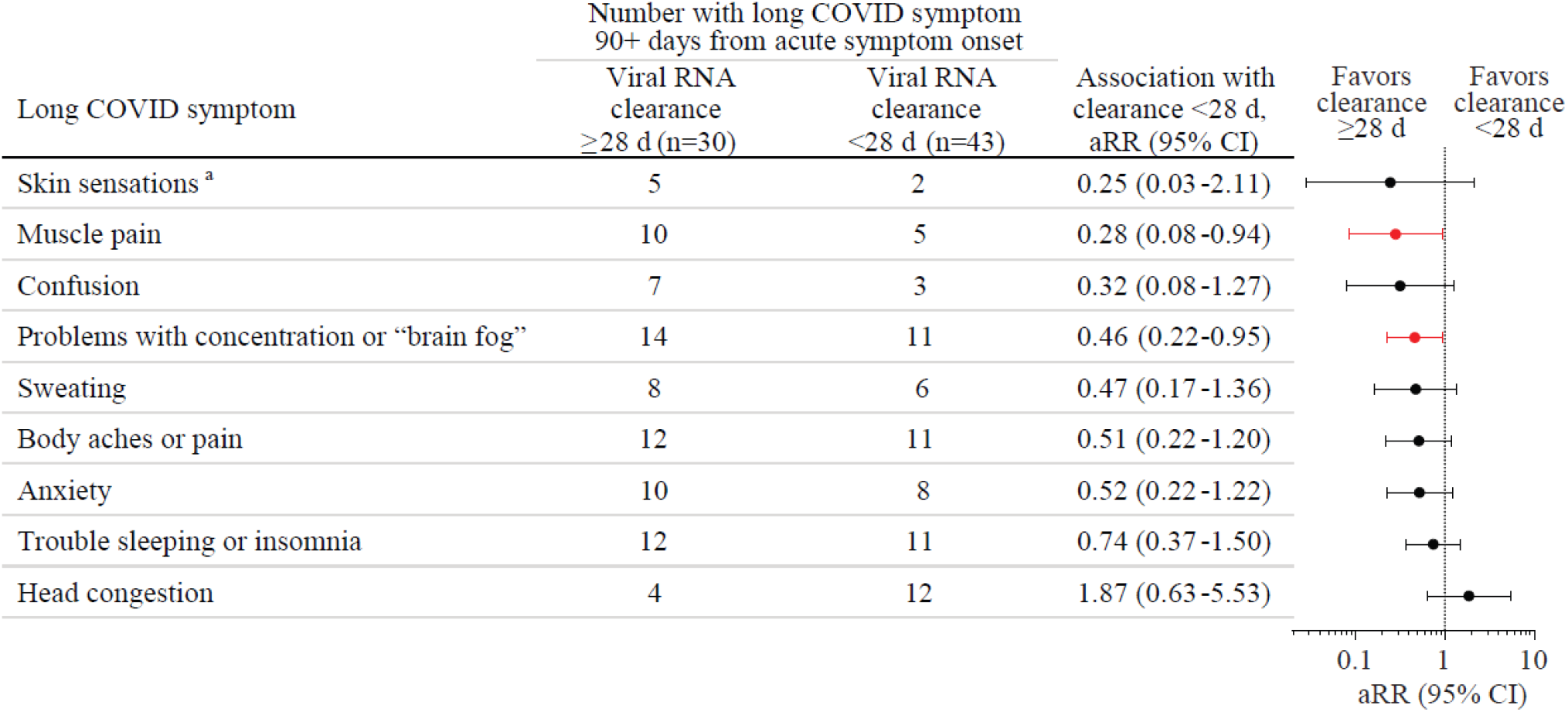
The association of the presence of each of 49 long COVID symptoms at 90+ days after acute COVID-19 illness onset with SARS-CoV-2 RNA clearance from the upper respiratory tract (URT, mid-turbinate nasal and/or saliva) within 28 days from acute COVID-19 onset was tested without adjustment using log-binomial regression analyses. Nine long COVID symptoms with p values <0.2 were tested using log-binomial regression analyses with adjustment for age, sex, BMI over/under 25, and COVID vaccination status prior to acute COVID-19 (1+ vaccines vs none), with results shown here. aRR, adjusted risk ratio, d, day. ^a^Tingling, numbness, coldness, or other unusual sensation in skin.

## Discussion

We demonstrate here that post-acute brain fog and muscle pain are associated with prolonged time to clearance of SARS-CoV-2 RNA from the upper respiratory tract during acute COVID-19. This finding provides evidence that delayed immune clearance of SARS-CoV-2 antigen and/or greater amount or duration of viral antigen burden in the upper respiratory tract during acute COVID-19 is directly linked to long COVID. This work indicates that host-pathogen interactions during the first few weeks after COVID-19 symptom have in impact on long COVID risk months later.

Currently, the biological mechanisms underlying long COVID and its myriad symptoms are unclear. There are several different hypotheses: persistent immune dysregulation following acute COVID-19, persistent SARS-CoV-2 virions or antigen (“reservoirs”) triggering chronic inflammation, reactivation of other viruses such as Epstein-Barr virus (EBV), autoimmunity triggered by acute illness, endothelial dysfunction and/or microthrombi impairing organ function, metabolic or mitochondrial dysfunction on the cellular level, autonomic nervous system injury, lingering or evolving organ injury from acute COVID-19, and microbiota alterations, among others (11). A recent comprehensive autopsy study provided evidence for an early viremic phase during acute COVID-19 that spreads infectious virus throughout the body and demonstrated convincingly that SARS-CoV-2 RNA can persist for >30 days in multiple extrapulmonary sites (12).

There have been a few studies that link long COVID with persistent SARS-CoV-2 antigen detected in the late post-acute time period, notably spike protein in plasma (13), SARS-CoV-2 proteins in plasma neuron- and astrocyte-derived extracellular vesicles (14), spike protein in monocytes (15), and fecal RNA shedding (16). However, there has been just one study to our knowledge linking virus kinetics or detection during acute COVID-19 with long COVID; this study associated the presence or absence of SARS-CoV-2 RNA in the plasma at the time of COVID-19 diagnosis and hospitalization with one or more symptoms of long COVID several months later (17). However, this study was limited to people with severe acute COVID-19 (17), and it may be that the presence of SARS-CoV-2 RNA-emia at hospital presentation is an indicator of the severity of acute COVID-19, which is already known to be a risk factor for long COVID (1).

Our study provides robust support to the hypothesis that viral kinetics during the acute phase of illness is associated with long COVID by using frequent longitudinal sampling beginning within days of acute COVID-19 onset to determine the duration of viral RNA shedding from the URT. Additionally, since long COVID may comprise one or more syndromes with unique mechanisms, we assessed individual symptoms from multiple organs or domains affected by long COVID to determine whether particular syndromes or symptom clusters of long COVID are associated with viral kinetics in the acute phase. We studied a population that primarily had mild-moderate COVID-19 and thus is more representative of the general population of people who had COVID-19 or long COVID and limits the amount by which severity of acute illness could confound our results. A final strength of this cohort is the racial and ethnic diversity of participants. A weakness of the study is that the intensity of study coordination involved in repeated longitudinal sampling and follow-up in acute and post-acute COVID-19 limited the number of participants enrolled.

Our data are consistent with a few biologically plausible scenarios. First, both delayed clearance of viral antigen and long COVID are caused by another factor such as dysregulated or distracted immunity. This dysregulated immunity might cause long COVID by reactivating other latent viruses, triggering autoimmunity, causing direct organ damage, or delaying tissue repair from acute infection. Second, persistent expression of viral antigens, and/or a larger burden of initial antigen, might directly cause long COVID by sustaining tissue injury and inflammation. The latter mechanism is supported by early studies linking antiviral treatment of people during acute COVID-19 with reduced incidence of long COVID (18). More studies with longitudinal repeated sampling beginning from acute COVID-19 illness treated with and without antivirals through the post-acute phase are urgently needed to assess how host-pathogen interactions lead to long COVID.

Here we report that at least two long COVID symptoms - brain fog and muscle pain – at 90+ days from acute COVID-19 onset are specifically associated with longer time to clearance of SARS-CoV-2 RNA from the upper respiratory tract during acute illness.

## Data Availability

All data produced in the present study are available upon reasonable request to the authors.

## Author Contributions

AA, TY, YM: conception and design. AA, TY, CH: statistical analysis and interpretation of data. PB, YM: obtained funding. DT, YM: administrative, technical, or material support. All authors: writing, review, and critical revision of the manuscript for important intellectual content. AA, YM: study supervision.

## OutSMART Study Group

M. Gabriela Varela Heslin, Sarika K. Mullapudi, Chamia Dorsey, Christine Payton, Sidney-Saint-Hilaire, Zihan Yang, Justin Chan, Razvan Azamfirei, Minyoung Jang, Taylor Church, Carolyn Reuland, Vismaya S. Bachu, Jennifer L. Townsend, Sara C. Keller, Jeanne C. Keruly, Justin P. Hardick, Madison Conte, and Thelio Sewell.

## Acknowledgments

We express gratitude for all the participants who donated time and samples for this study. We thank the following individuals for their assistance with this study: Diane M. Brown, Brittany Barnaba, Curtisha Charles, Michelle Prizzi, Oyinkansola Kusemiju, and Jaylynn R. Johnstone.

